# The iPSYCH2015 Case-Cohort sample: updated directions for unravelling genetic and environmental architectures of severe mental disorders

**DOI:** 10.1101/2020.11.30.20237768

**Authors:** Jonas Bybjerg-Grauholm, Carsten Bøcker Pedersen, Marie Bækvad-Hansen, Marianne Giørtz Pedersen, Dea Adamsen, Christine Søholm Hansen, Esben Agerbo, Jakob Grove, Thomas Damm Als, Andrew Joseph Schork, Alfonso Buil, Ole Mors, Merete Nordentoft, Thomas Werge, Anders Dupont Børglum, David Michael Hougaard, Preben Bo Mortensen

## Abstract

The Lundbeck Foundation Integrative Psychiatric Research (iPSYCH) consortium has almost doubled its Danish population-based Case–Cohort sample (iPSYCH2012). The newly updated cohort, named iPSYCH2015, expands the study base with 56,233 samples, to a combined total of 141,265 samples. The cohort is nested within the Danish population born between 1981 and 2008 and is a Case-Cohort design including 50,615 population controls. We added more cases to the existing phenotypes identified with, schizophrenia (N_new_=4,131/N_total_=8,113), autism (N_new_=8,056 / N_total_=24,975), attention-deficit/hyperactivity disorder (N_new_=10,026/N_total_=29,668) and affective disorder (N_new_=13,999/N_total_=40,482) of which a subset has bipolar affective disorder (N-_new_=1,656/N_total_=3,819). We also added two additional focus phenotypes, schizophrenia spectrum disorder (N=16,008) and post-partum disorder (N=3,421). In total, the expanded iPSYCH2015 sample consists of 93,608 unique individuals in the case groups and 50,615 population controls. For the sample expansion, DNA was extracted and amplified from dried blood spots samples stored within the Danish Neonatal Screening Biobank and genotyped using the Illumina Global Screening Array. The Biobank sample retrieval rate was 95%, and the genotyping success rate was 92% (97% of retrieved). We expanded the follow-up period by three years, including data such as longitudinal information on health, prescribed medicine, social and socioeconomic information.

## Introduction

Psychiatric disorders (PD), such as schizophrenia and autism, are common and highly heritable. Prior reports have estimated that one-third of the population in the middle- and high-income countries will be affected by at least one PD during their lifespan^1^. PD can be seriously disabling and leads to numerous adverse outcomes, such as lower educational attainment^2^. Patients with PD sometimes fall behind their contemporaries, both at a social and economic level^3–6^. PDs also leads to physical-health complications^7^, and those affected by PD face a more than 2-fold increased mortality rate ratio^8–10^. Beyond the ramifications, at the personal level, PD also has severe effects on society. In 2010, the EU had 165 million people affected by PD, and PD was the leading cost of disability in any disease group^11–13^.

In recent years, great strides have been made towards an understanding of the aetiology of psychiatric disorders. Despite the high heritability of PD, the phenotype derives from many small non-deterministic genetic and environmental contributions. This complicates the identification of the individual components and complicates assessment of individual impacts on the phenotype.^14^

Many cohorts used to study diseases, including PD, are samples of convenience. While these cohorts have values in the study of biology, it is well documented that eg. the UK Biobank participants are on average older than non-participants, and they are more likely to be female, to better off socioeconomically, to be less obese, to not smoke or drink alcohol and overall, they report fewer health conditions^15^. The skewed ascertainment may result in biased estimates and incorrect inference downstream since the sample does not represent the source population. This can result in recommendations and suggested interventions being misleading or even invalid^16^.

The population-based case-cohort sample design of the Lundbeck Foundations Initiative for Integrative Psychiatric Research (iPSYCH) is recruited as to avoid ascertainment bias and thus to reflect the underlining population. iPSYCH selected its Case-Cohort design from the population, and we were thus enables unbiased estimation absolute risks and incidence rates. The random population base makes it possible to estimate the effect sizes of genetic markers, which are representative of the entire Danish population. The iPSYCH2012 cohort is a unique resource made possible by leveraging the all-encompassing Danish electronic health records in combination with population-wide Biobanks.^17^

The iPSYCH2012 cohort was used in the first population-based study of cross-disorder interactions in PD, which suggested that dysregulation during neurodevelopment plays a general role^18^. Through Genome-Wide Association Studies (GWAS) iPSYCH2012 data was central in discovering the first 12 loci for attention-deficit/hyperactivity disorder (ADHD)^19^, as well as the first five common variant loci for Autism Spectrum disorder (ASD)^20^. Additional to ASD and ADHD, the primary diagnosis of the iPSYCH consortia is Bipolar Disorder (BD), Affective Disorder (AD) and Schizophrenia (SCZ). Through the Psychiatric Genetics Consortium(PGC)-iPSYCH collaboration, the data were used for landmark papers in all primary disorders of the iPSYCH^21–26^.

Complementary to the genetic discovery, genetic liability can be aggregate through additive models such as Polygenic Risk Scores(PRS). When combined with the Danish registries PRS becomes a powerful tool for studying interactions between genetic and other risk factors (e.g. environmental variables). We applied this model to the study of interaction one or more PD with the psychosocial environment^27^, Nitrogen Dioxide^28^, Unipolar Depression in Early Life^29^, substance use disorders^30^, voter turnout^31^ and Parental Socioeconomic Status coupled with history of PD^32^

Besides being ideal for studying the selection diagnosis, the population-based design enable studies of phenotypes not considered in the initial design through linkage with registries^17^. Investigators have used the iPSYCH2012 cohort to study adolescent residential mobility^33^, childhood asthma^34^, penetrance of PD in 22q11 syndrome^35^, the effects of the mitochondria^36,37^, anxiety^38^, suicide^39^ and cannabis use disorder^40^.

In addition to standard genome-wide-association studies of psychiatric disorders the iPSYCH samples are used for multiple other types of markers. Nested subsets of the iPSYCH samples were used to study intermediate molecular phenotypes. We investigated how methylation might mediate PD in 22q11 carriers^41^. We integrated genetic and epigenetic data to investigate ASD and found the PRS predicted methylation changes^42^. Finally, we investigated if bloodborne neurotropic and inflammation markers are associated with psychiatric diseases, and found associations with Brain-derived neurotrophic factor(BDNF) levels in ASD^43^.

While the iPSYCH2012 cohort has yielded significant results and has contributed to landmark papers in the study of PD, it still has the potential to contribute significantly to science in years to come, the value of iPSYCH is scalable by increasing the number of samples included. This perspective describes the update of the existing iPSYCH2012 to the iPSYCH2015 Case-Cohort study. We reiterate the core epidemiological design and describes the updated analytical approach. With this update, the total number of samples in the Case-Cohort is 141,265 of which 129,950 have been genotyped. Simultaneously we expand the recruitment with three more birth years, by including all diagnosis up to 2015 for the entire sample.

### Design

The iPSYCH2015 Case-Cohort expands on the design used in the iPSYCH2012 version^17^. Recruitment is done through the use of unique Central Person Register (CPR) numbers. All residents in Denmark are assigned a CPR-number either upon birth or at the time of immigration to Denmark. CPR is linked to the national population-wide registries (e.g. Danish Psychiatric Central Research registry) to recruit participants^17^. We cross-referenced CPR with the availability of dried blood spot (DBS) in the Danish Neonatal Screening Biobank (DNSB). The DNSB has been storing residual DBS since May 1981 and encompasses close to all births in Denmark since then. While the DNSB is a screening Biobank with primary activities centred around new-born screening, ancillary and secondary purposes, including research, are allowed provided that relevant authorities have granted approval^44^.

Once the initial selection and data generation is completed, the CPR-number allows continuous updates of information from population-based registers through pseudonymization.

### Cohort-nomenclature

We here present an addition, which is complementary to our prior cohort. The two parts can be referred individually or combined. iPSYCH2012 refers exclusively to the original selection, which we accounted for in our previous perspective^17^. iPSYCH2015i refers solely to the second selection, which we account for within this perspective. iPSYCH2015 refers to the combined iPSYCH2012 and iPSYCH2015i

### Expanding the study base

The study base for iPSYCH2015i includes all singleton births to mothers who are living in Denmark (i.e. mother has a CPR-number) between 1st of May 1981 and 31st of December 2008, where the child was alive and resided in Denmark at their one-year birthday (N=1,657,449). The study base from iPSYCH2012 (1981-2005) is expanded with individuals born from 2006 to 2008.

Samples in the population-based cohort were selected randomly from the study base. In iPSYCH2012 30,000 individuals born between 1st of May 1981 and 31st of December 2005 were randomly selected and in iPSYCH2015i 21,000 individuals born between 1st of May 1981 and 31st of December 2008 were randomly selected. Due to the random selection, 385 samples were selected for both iPSYCH2012 and iPSYCH2015i. As a consequence of the selection criteria, 2958 individuals in the population-based cohort had at least one of the focus disorders in iPSYCH2015.

The probability of being selected for the population-based cohort depends on the year of birth as only the years 1981-2005 are included in both selections. The probability of being selected if born between 1st of May 1981 and 31st of December 2005 is 3.27% (48,227/1,472,808). The probability of being selected for those born between 1st of January 2006 and 31st of December 2008 is 1.29% (2,388/184,641). A breakdown samples include per year in the population-based cohort is shown in Supplementary table 1.

### Diagnosis of Mental Disorder

PD diagnosis was identified by linking relevant registries to the study base; we outlined this in details in the iPSYCH2012 perspective^17^, but a brief summation follows. Psychiatric diagnoses until the 31st of December 2015 were used, which added three years of follow up compared with iPSYCH2012.

For ASD (F84.0, F84.1, F84.5, F84.8 or F84.9), ADHD (F90.0), BP (F30-F31) and AD (F30-F39), no changes were made in the inclusion criteria. Besides Schizophrenia (F20), the whole Schizophrenia spectrum (F20-F29) was included. Finally, we added post-partum depression as a diagnosis defined as any psychiatric disorder (F00-F99) within one year of giving birth to a living child.

In total, 36,741 additional cases were identified across all phenotypes bringing the total number of participants affected by PD to 93,608. Detailed numbers for the iPSYCH2015 cohort are shown in table 1. The numbers added with the iPSYCH2015i is shown in supplementary table 2.

**Table 1:** Number of persons included in iPSYCH2015 combined population-based sample of the Danish population born 1981-2008.

| Group membership<sup>a</sup> | ICD10 diagnoses<sup>b</sup> | Follow-up period | Number of persons |  |  |
| --- | --- | --- | --- | --- | --- |
|  |  |  | Initial Sample | Samples genotyped | Passed Sample QC |
| Schizophrenia | F20 | 1994-2015 | 8113 | 7538 (93%) | 7323 (90%) |
| Schizophrenia spectrum disorder | F20-F29 | 1994-2015 | 16008 | 14930 (93%) | 14480 (90%) |
| Bipolar disorder | F30-F31 | 1994-2015 | 3819 | 3577 (94%) | 3461 (91%) |
| Affective disorder | F30-F39 | 1994-2015 | 40482 | 37996 (94%) | 36823 (91%) |
| Autism | F84.0, F84.1, F84.5, F84.8 or F84.9 | 1994-2015 | 24975 | 24095 (96%) | 23218 (93%) |
| ADHD | F90.0 | 1994-2015 | 29668 | 28508 (96%) | 27562 (93%) |
| Post partum disorder | F00-F99<sup>c</sup> | 1994-2015 | 3421 | 3103 (91%) | 2974 (87%) |
| Any case | All ICD codes listed above | 1994-2015 | 93608 | 88831 (95%) | 85891 (92%) |
| Population-based cohort | Random sampling, i.e. disregarding any diagnostic information |  | 50615 | 48420 (96%) | 46760 (92%) |
| Total number of persons |  |  | 141265 | 134455 (95%) | 129950 (92%) |

Abbreviations: ADHD, attention-deficit/hyperactivity disorder; ICD-10, International Classification of Diseases, 10th revision; iPSYCH, Integrative Psychiatric Research; QC, quality control, <sup>a</sup>Groups are not mutually exclusive. <sup>b</sup>Initial ICD10 diagnosis used to select case groups. Identification was performed through linkage to the Danish Psychiatric Central Research Register. <sup>c</sup>Psychiatric diagnosis within one year after giving birth to a live-born baby. Percentages is relative to the initial sample of the same group membership.

It should be noted that Table 1 and Supplementary Table 2 account for 1000 persons in the population cohort selected after the criteria described above for the iPSYCH2015i population cohort; however, they were part of a separate anorexia study. These data are not accounted for in the following sections on retrieval and processing.

### The Danish Neonatal Screening Biobank

Once identified, residual DBS from the Neonatal Screening efforts in Denmark were retrieved from the DNSB. Hollegaard *et al*. (2009) have described the processing of samples in detail, and protocol modifications are described in Bækvad-Hansen *et al*. (2017)^45,46^. Experimentally, the processing of the iPSYCH2015i sample is identical to that used in iPSYCH2012; however, we made minor modifications to eliminate bottlenecks and increase laboratory efficiency.

An overview of the pipeline used to process the samples is shown in figure 1.

**Figure 1.**
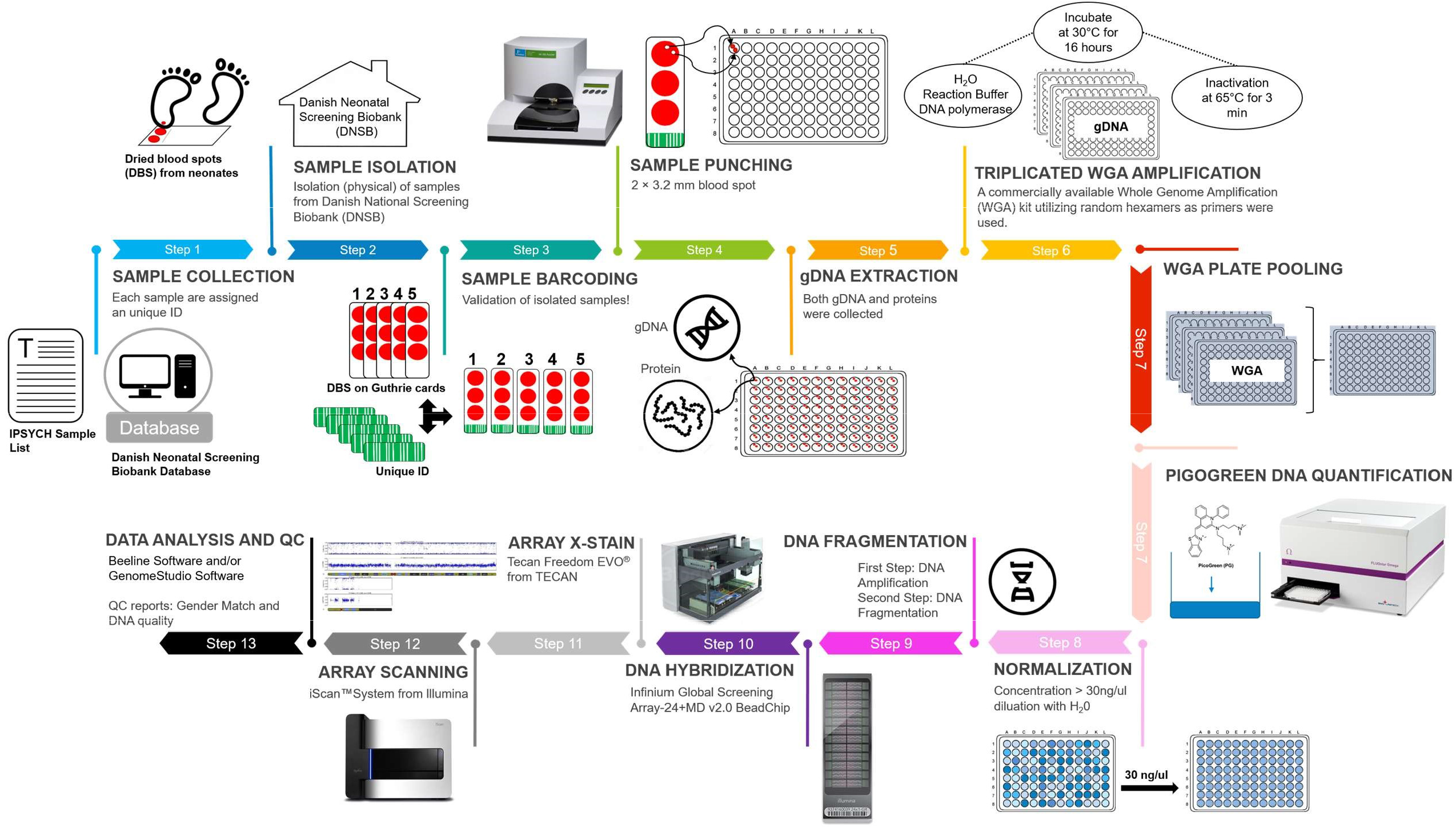
Overview of the pipeline starting with the CPR-number and ending with genotype calling. Briefly, using the CPR-number as a starting point, the biological material is retrieved and assigned a unique pseudonymization ID. The protein fraction is extracted and kept for future analysis. Genomic DNA is then extracted and whole genome amplified in triplicates. The triplicates are then pooled and concentrations are measured. The samples are normalized and finally randomized across new plates. The samples are then processed using the Illumina GSA v2 + MD array, the genotypes are trained, then called and samples quality and integrity are verified.

In the following, we will account for the pipeline and explicitly state the difference between the processing of iPSYCH2012 and iPSYCH2015i.

Once linked through the CPR-number, the DBS cards were retrieved from storage and then given a unique pseudonymization identifier. Each sample had two 3.2mm DBS spots excised. Proteins were extracted and stored. DNA was extracted and Whole Genome Amplified (WGA) in triplicates. Where iPSYCH2012 used PCR machines for isothermal WGA, iPSYCH2015i used an incubation-oven to increase daily throughput by several folds.

Following WGA and pooling of the triplicates, DNA concentrations were measured. For iPSYCH2012, we excluded samples with DNA concentrations below 15 ng/µL. In iPSYCH2015i, we attempted to genotype all samples. If possible, we diluted to 30ng/µL; otherwise, we processed the samples as they were.

While DNA concentration is a predictor of performance, more than 90% of samples with concentrations below 15ng/µL still pass a call rate threshold of 95%. A more detailed breakdown of the effects of low concentration samples is available in supplementary figure 1.

After determination of concentrations and dilution, the samples were randomized onto new plates, before being genotyped using the Global Screening Array v2 with a Multi disease drop-in. Array choice is a modification from iPSYCH2012 where genotyping was done using the PsychArray V1.0. Genotyping was done according to the manufacture’s instruction with the modification that the “Make MSA” step was carried out using 7µL sample at 30 ng/µL and 1µL 0.4N sodium hydroxide.

While a change of array could be considered an incompatibility, robust statistical tools exist to address this challenge. Wrey *et al*. (2018) exemplify this; here, 33 cohorts are meta-analysed across 13 different countries using 11 different arrays resulting in 44 risk loci for major depressive disorder^22^. While the change of array should be taken into account in analysis (e.g by analysing iPSYCH2012 and iPSYCH2015i separately followed by meta-analysis), a valid conclusion can still be drawn.

Once scanned, the genotypes were called based on a custom cluster trained on the first 4,158 samples. Further details on training variant call are shown in supplementary text 1. Call rates were determined for each sample, and samples with call rates 95% were excluded. Sex was estimated based on heterozygosity of the X-chromosome for samples passing the call rate threshold. The estimated sex was compared to that recorded in the medical birth registry, and divergent calls were manually verified based on log R ratio and B Allele frequency for chromosomes X and Y^47^.

### Sample availability and genotyping

We identified 55,530 cases and controls matching the criteria for inclusion. We compared these against the samples database at the DNSB. We were unable to locate 1.6% of the selected samples within the DNSB. This loss is due to a combination of parents electing not to screen their children, and that the age of the Biobank predates the advent of electronic records leading to gaps and inconsistencies.

A further 2.9% of samples were in the records, but could not be located physically in the biobank. Finally, about 0.1% of the samples were retrieved, but did not have sufficient material remaining for inclusion in the study as per the priorities of the DNSB^44^. A detailed overview of sample drop out is shown in supplementary figure 2.

**Figure 2.**
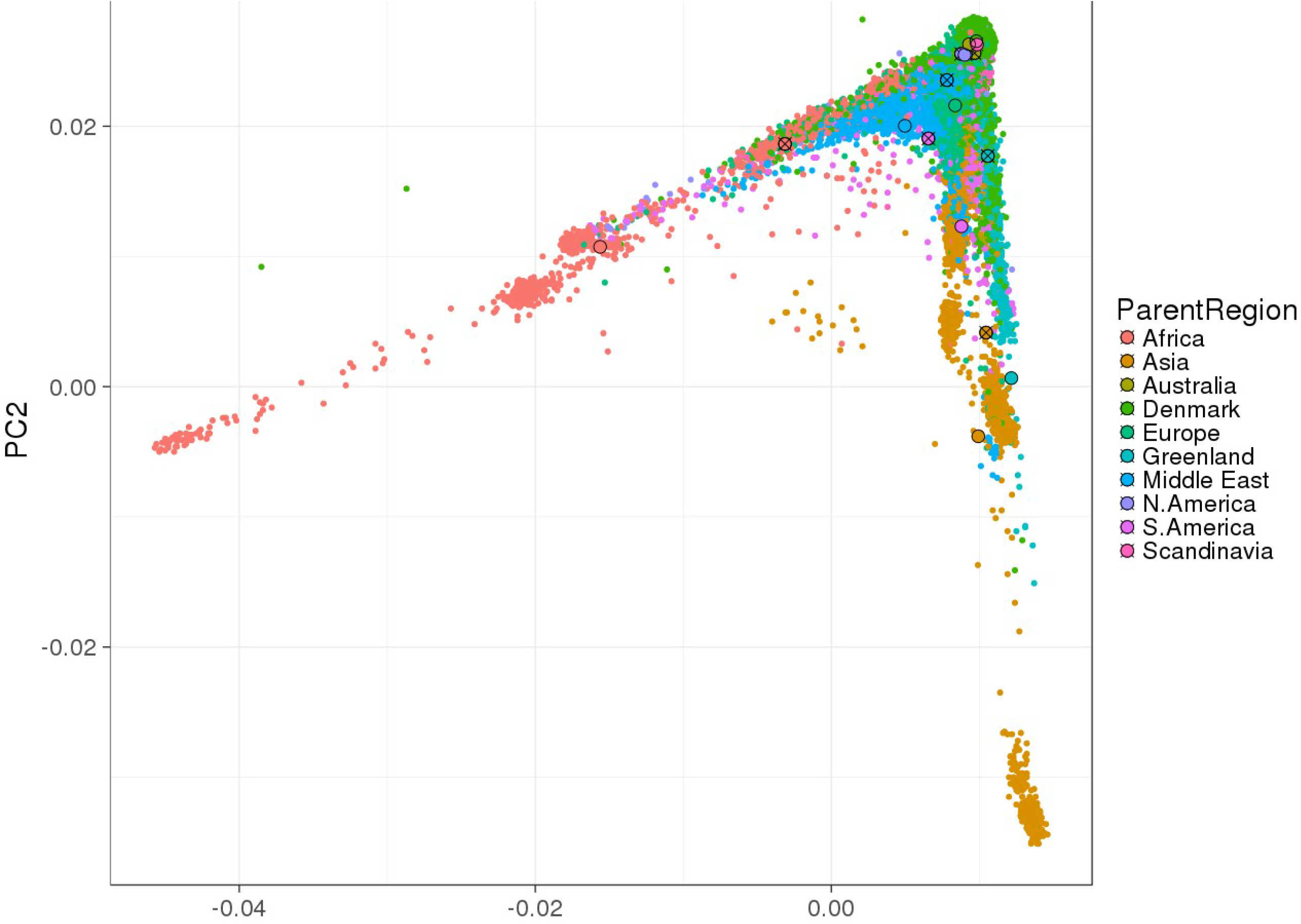
Scatterplot of the first two principal components coloured according to the parental region of birth. Values are calculated using all 130 thousand iPSYCH samples (iPSYCH2012 plus iPSYCH2015i). Circles colour indicate mean values for the given parental group. Crossed Circles indicates both parents born abroad (region indicated by the colour). Non-crossed circles indicate one Danish-born parent and one parent born in the region indicated by the colour. Majority of samples and thus parental origins are clustered in the densest part of the PCA plot, while hard to distinguish the implications are that these subsets within the cohort are heterogenous.

In total, we located 95.5% of the selected samples and genotyped them. First, we evaluated samples according to their percentage of called genotypes (call rate), which we required to be higher than 95%, 2.3% of the samples failed to meet this criteria. Secondly, we verified if a sex call based on genotype matched that expected that recorded in the birth registry. We evaluated all samples with mismatches between estimated and recoded sex using SNP array-based karyotyping. We included the samples that filed the sex check if there existed a valid reason (e.g. Aneuploidy) or the registry was ambiguous (e.g. gender change). Samples failing the sex check without a valid reason was dropped from the study. A plot stratifying on genotype performance and sex estimates are shown in supplementary figure 3. A summary of reasons for sex mismatches is shown in supplementary figure 4.

### Population heterogeneity

To visualize population structure and heterogeneity, we conducted a principal component analysis of all 129,950 samples with genotyping in iPSYCH2015. Detailed description of calculation for the PCA plot is shown in supplementary text 2. The first two principal components separated samples along continental vectors mirroring geographical distance, see figure 2. The PCA plots for iPSYCH2012 and iPSYCH2015i are shown in supplementary figure 5.

Similar to iPSYCH2012, visual inspection indicates that parental place of birth from the registries predicted clustering. When both parents were of Danish or Scandinavian origin, they clustered in the densest part of the PCA plot. The individuals with foreign parents were genetically most distant from the densest center of the PCA plot, and those with one Danish and one foreign were intermediate.

We defined the centre of an ellipsoid based on the mean value for principle components one through three. We used data from only those participant where registries allowed us to trace their genealogy back to four grandparents born in Denmark. We then calculated the standard deviation and defined the length of each axis in the ellipsoid, as five standard deviations along the relevant axis. The resulting ellipsoid encompasses 91.3% of all samples in the cohort, confirming that the Danish population is primarily homogenous European by descent.

### Ethical framework

The Danish Scientific Ethics Committee, the Danish Health Data Authority, the Danish data protection agency, the Danish Neonatal Screening Biobank steering committee and Statistics Denmark, approved this study. The requirement of multiple permissions is in keeping with the ethical framework and the Danish legislation protecting the use of these samples. ^44,48,49^

Permission has been granted to study genetic and environmental factors for the development and prognosis of mental disorders. To unravel the foundation of severe mental disorders, it is central that this rich data source is accessible to the international research community to the largest extent possible. It is also paramount to protect the privacy of the individuals included in the study. Due to the sensitive nature, data can only be accessed through secure servers where export of sensitive information is prohibited and prevented. For this, the iPSYCH consortia have created a framework and data access model; please contact the authors for requests on collaboration.

Throughout the entire study, authorized researchers may access pseudonymised data only. The link to the CPR-number is stored in a separated and secure environment accessible to restricted personnel only. This link allows us to update and refine phenotypes if granted by relevant authorities.

### Perspectives

With the iPSYCH2012 cohort, we created a foundation of 77,639 genotyped samples that have contributed significantly to a large number of milestone papers in psychiatric genetics^18–24^. iPSYCH2015 includes iPSYCH2012 and is an expansion both in numbers of cases, numbers of controls as well as years of follow-up. With the new data in the iPSYCH2015, we have added 52,301 samples expanding the total cohort to 129,950 successfully genotyped samples. The study includes detailed phenotypic information on relevant PD, relevant health information, as well as social factors.

The perspective for the expanded cohort is immense. Not only does it offer the largest known unbiased population-based cohort, but it also provides a more than 90% participation rate of all known cases with PD in Denmark. With the new data, researchers will have access to a resource that will facilitate discovery in psychiatric genetics for years to come. Plans for studies on the expanded iPSYCH2015 cohort are numerous, and in the following, we present a small subset of the analysis planned.

iPSYCH2012 made significant contributions in GWAS into PD. The expansion into iPSYCH2015 will add 36,741 cases, which add to the statistical power and thus higher potential for discovery. The second cohort also opens the possibility of doing discovery and validation on two cohorts nested within an overlapping and almost identical baseline population.

New iterations of the GWAS studies are underway in collaboration with the PGC and other international partners. Collaborations exist both at the level of the individual diagnosis, but also examining comorbidities and general risk (cross disorder). We expect that this will increase the knowledge about the non-deterministic genetic components that contribute to the development of PD. The knowledge gained will increase the potential of derived analysis. One example of a type of research that will benefit are those that leverage polygenic risk scores and registry information, that aim to delineate the complex multifactorial contributions that might be at the core of risk for PD.

An essential part of the next generation of analysis is to understand further how differences in DNA translates into a disorder at the organism level. Another possibility is to explore intermediate molecular phenotypes. This hypothesis will be tested in several ways; one example is using quantitative-trait-loci based estimators of gene expression to test for altered gene expressions in brain tissue^50^.

A more direct approach bridging the gap between genotype and phenotype is to directly measure the intermediate molecular phenotypes in nested subsets of the cohort. One sub-project will add immunohistochemically measures of neurotrophic factors in 30 thousand samples focusing on ASD and ADHD. Researchers within iPSYCH will perform metabolomics on the polar fraction in approximately 10 thousand samples again with a particular focus on ASD and ADHD. There is also an ongoing project to measuring RNA-seq on 580 monozygotic twin pairs of which most are discordant for PD.

Common for all the molecular phenotypes is that we are limited to measurements that are detectable in blood. While blood is not the primary tissue of interest for disorders of the central nervous system, blood still has the potential to yield knowledge about diagnostic or prognostic markers, which could significantly aid in future diagnostic applications.

The aggregation of various measures on a highly overlapping sample allows for multi-layered multiomics analysis with unique opportunities for discoveries. With our combinatory studies, we have a unique opportunity to discover if trajectories towards PD is already established at birth.

The research just described will contribute essentially to understanding the aetiology of PD. However, it is also vital that we go beyond the biological understanding and attempt to operationalize the knowledge gained. One example of such is a project which uses pharmacogenetic to explore low efficacy of pharmaceutical treatment options for PD.^51^

Only through rigorous work requiring detailed phenotypes, coupled with reliable molecular measures of predictive value, can we hope to bring about the promise of precision medicine in psychiatry, with the iPSYCH2015 case-cohort we have added an essential resource towards realising that promise.

## Data Availability

Data availability is limited due to the sensitive nature. We are working on GDPR compliant models for remote access; please contact the authors for more details.

## Acknowledgements

The Authors would like to thank Vivek Appadurai for the base calculations for the PCA plot. This study was supported by The Lundbeck Foundation, the Stanley Medical Research Institute, an Advanced Grant from the European Research Council (project number 294838) and Centre for Integrated Register based Research at Aarhus University. This research has been conducted using the Danish National Biobank resource, supported by the Novo Nordisk Foundation.

---

**Supplementary Table 1:** Counts and observed sample frequencies among individuals selected for inclusion in the population-based cohort: Singletons born in Denmark 1st May 1981 to 31st December 2008, known mother, alive and resident in Denmark at the one-year birthday.

| Birth year | Number of males in study base | Number of males in population-based cohort | Frequency in percent | Number of females in study base | Number of females in population-based cohort | Frequency in percent |
| --- | --- | --- | --- | --- | --- | --- |
| 1981 | 17394 | 552 | 3.17 | 16811 | 589 | 3.50 |
| 1982 | 26150 | 799 | 3.06 | 24801 | 798 | 3.22 |
| 1983 | 25112 | 831 | 3.31 | 24058 | 790 | 3.28 |
| 1984 | 25655 | 835 | 3.25 | 24400 | 859 | 3.52 |
| 1985 | 26535 | 939 | 3.54 | 25402 | 873 | 3.44 |
| 1986 | 27487 | 896 | 3.26 | 26016 | 889 | 3.42 |
| 1987 | 28022 | 897 | 3.20 | 26270 | 857 | 3.26 |
| 1988 | 29313 | 922 | 3.15 | 27452 | 896 | 3.26 |
| 1989 | 30389 | 1014 | 3.34 | 28807 | 923 | 3.20 |
| 1990 | 31485 | 1066 | 3.39 | 29798 | 1013 | 3.40 |
| 1991 | 31810 | 1014 | 3.19 | 30149 | 999 | 3.31 |
| 1992 | 33559 | 1106 | 3.30 | 31637 | 1011 | 3.20 |
| 1993 | 33154 | 1007 | 3.04 | 31490 | 1052 | 3.34 |
| 1994 | 34164 | 1163 | 3.40 | 32629 | 1041 | 3.19 |
| 1995 | 34567 | 1099 | 3.18 | 32601 | 1063 | 3.26 |
| 1996 | 33269 | 1032 | 3.10 | 31366 | 968 | 3.09 |
| 1997 | 33149 | 1063 | 3.21 | 31403 | 1062 | 3.38 |
| 1998 | 32386 | 1063 | 3.28 | 30607 | 974 | 3.18 |
| 1999 | 32311 | 1014 | 3.14 | 30778 | 1020 | 3.31 |
| 2000 | 32810 | 1087 | 3.31 | 31085 | 1063 | 3.42 |
| 2001 | 31741 | 1024 | 3.23 | 30339 | 1017 | 3.35 |
| 2002 | 31327 | 1067 | 3.41 | 29518 | 942 | 3.19 |
| 2003 | 31479 | 1019 | 3.24 | 29894 | 1006 | 3.37 |
| 2004 | 31383 | 1018 | 3.24 | 29926 | 974 | 3.25 |
| 2005 | 31134 | 996 | 3.20 | 29786 | 1025 | 3.44 |
| 2006 | 31761 | 393 | 1.24 | 29992 | 403 | 1.34 |
| 2007 | 31229 | 386 | 1.24 | 29788 | 401 | 1.35 |
| 2008 | 31859 | 415 | 1.30 | 30012 | 390 | 1.30 |
| Total | 850634 | 25717 |  | 806815 | 24898 |  |

By design we aimed 30,000 randomly selected individuals born 1981-2005 and 21,000 born 1981-2008 (50,615 in total) from the population-based cohort (N = 1,657,449). The probability of being selected for the population-based cohort depends on the year of birth as only the years 1981-2005 are included in both selections. The designed probability of being selected if born between 1st of May 1981 and 31st of December 2005 is 3.27% (48,227/1,472,808). The designed probability of being selected for those born between 1st of January 2006 and 31st of December 2008 is 1.29% (2,388/184,641).

**Supplementary Table 2:** Number of persons included in iPSYCH2015i population-based sample of the Danish population born 1981-2008.

| Group membership<sup>a</sup> | ICD10 diagnoses<sup>b</sup> | Follow-up period | Number of persons |  |  |
| --- | --- | --- | --- | --- | --- |
|  |  |  | Initial Sample | Samples genotyped | Passed Sample QC |
| Schizophrenia | F20 | 1994-2015 | 4131 | 3996 (97%) | 3910 (95%) |
| Schizophrenia spectrum disorder | F20-F29 | 1994-2015 | 9111 | 8661 (95%) | 8419 (92%) |
| Bipolar disorder | F30-F31 | 1994-2015 | 1656 | 1578 (95%) | 1532 (93%) |
| Affective disorder | F30-F39 | 1994-2015 | 13999 | 13339 (95%) | 12992 (93%) |
| Autism | F84.0, F84.1, F84.5, F84.8 or F84.9 | 1994-2015 | 8056 | 7799 (97%) | 7650 (95%) |
| ADHD | F90.0 | 1994-2015 | 10026 | 9641 (96%) | 9420 (94%) |
| Post partum disorder | F00-F99<sup>c</sup> | 1994-2015 | 1890 | 1734 (92%) | 1641 (87%) |
| Any case | All ICD codes listed above | 1994-2015 | 36741 | 35069 (95%) | 34156 (93%) |
| Population-based cohort | Random sampling, i.e. disregarding any diagnostic information |  | 19982 | 19086 (96%) | 18597 (93%) |
| Total number of persons |  |  | 56233 | 53688 (95%) | 52301 (93%) |

Abbreviations: ADHD, attention-deficit/hyperactivity disorder; ICD-10, International Classification of Diseases, 10th revision; iPSYCH, Integrative Psychiatric Research; QC, quality control, <sup>a</sup>Groups are not mutually exclusive. <sup>b</sup>Initial ICD10 diagnosis used to select case groups. Identification was performed through linkage to the Danish Psychiatric Central Research Register. <sup>c</sup>Psychiatric diagnosis within one year after giving birth to a live-born baby. Percentages is relative to the initial sample of the same group membership.

