## Supplementary information for "The iPSYCH2015 Case-Cohort sample: updated directions for unravelling genetic and environmental architectures of severe mental disorders"

### Supplementary Text 1. Training variant calls

The first 4,158 samples processed were used to train the variant caller (cluster positions), which defines all genotype calls in the iPSYCH2015i cohort. Samples were roughly quality controlled using a generic cluster file^1^; samples were removed if call rates were below 97% or the estimated sex differed from electronics records. Genotype call training was done using GenTrain V3. First, autosomal variants were trained. Next variant on the X-chromosome was trained using only Female Samples, and finally, Y-chromosomes were trained using only male samples.

Once genotype training was completed, the loci were manually inspected if:

1. Call frequency was below 0.9 (less than 90% of samples called in given loci),
2. Cluster separation was below 0.15. (indicating poor separation of variants)
3. Heterozygosity Rate >50% (indicating bad clustering or annotation of variant)
4. Heterozygosity on chromosome Y
5. Heterozygosity in males on chromosome X

During inspection variant, calls were adjusted or excluded as appropriate. While the training set was curated for the above conditions, they may still be present in the final call set as new samples might deviate from the stated conditions. Following completion, the cluster positions were exported and used to QC the entire cohort and formed the basis of the final variant call sets.

**References**

1.	Infinium Global Screening Array v2.0 Product Files. https://support.illumina.com/downloads/infinium-global-screening-array-v2-0-product-files.html.

### Supplementary Text 2. Principal Component Analysis (PCA)

The principal components were calculated relative to the 1000 genomes project phase 3^1^ call sets(1KGP) using PLINK^2^. Initially, variants with a Minor Allele Frequency of <5% were removed from 1KGP. Variants with Hardy-Weinberg Equilibrius P < 10E-6 were removed. The remaining variants were LD pruned removing r^2^ values >0.1 in 1kb windows. Finally, the 1KGP data were merged with iPSYCH2015 leaving 171,547 variants for the PCA calculations.

The PC’s were plotted using R and GGplot2. Samples were coloured based on parental regions of birth. If both parents were born in Denmark, it was coloured as such, and else the sample was coloured by the foreign parents country.

2.	Chang, C. C. et al. Second-generation PLINK: rising to the challenge of larger and richer datasets.

### Supplementary Figures

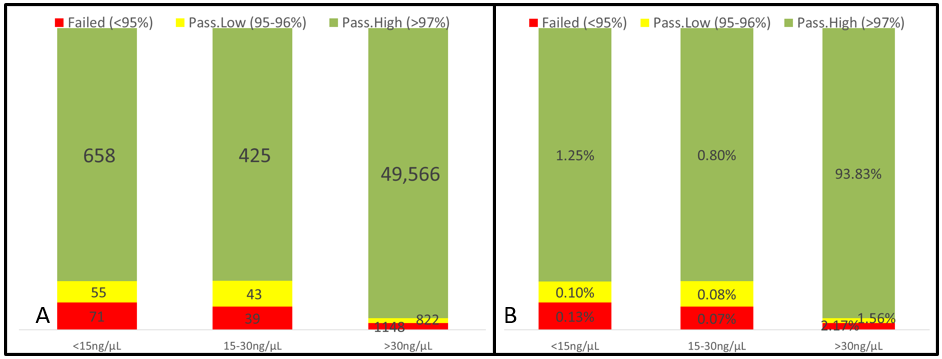

Supplementary Figure 1. Where iPSYCH2012 dropped all samples with concentrations below 15 ng/µL before genotyping, iPSYCH2015i attempted to genotype all samples regardless of concentration. While 2% of the samples in iPSYCH2015i had concentrations below 15ng/µL, most low concentration samples resulted in call rates above 97%. We observe the same performance for concentrations in the range 15-30ng/µL as we do below 15ng/µL, indicating that the previously used threshold was entirely arbitrary. While concentrations above 30ng/µL results in a larger number of high-quality genotyping, more than 90% of samples in any bin is considered of high quality. Given the success rate of low quantity samples relative to the cost of creating a cohort, we recommend attempting all genotypes regardless. These data do not reflect if the genotype sex calls match the expected, and only considers the effects of low concentrations samples on Genotyping call rates.

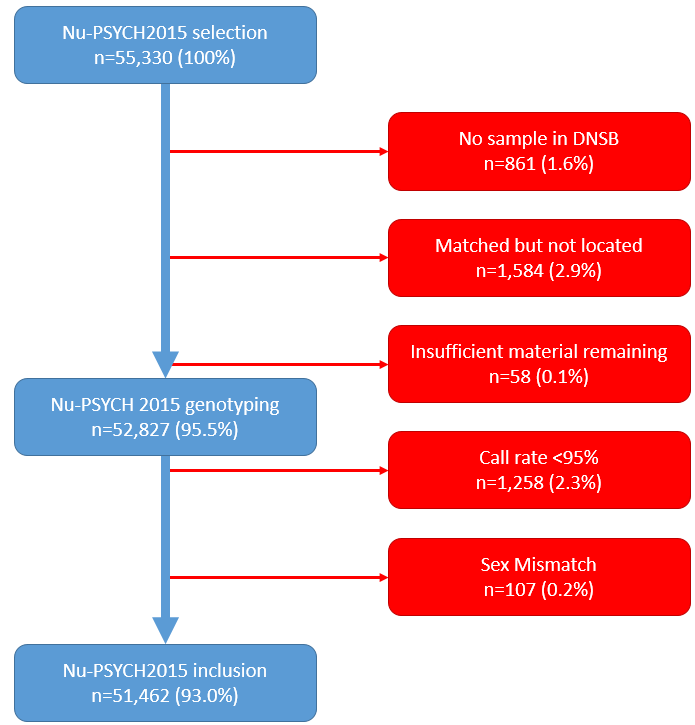

Supplementary figure 2. A detailed outline of samples dropout, accounting for all dropped samples starting with the selection until the pass samples QC genotypes.

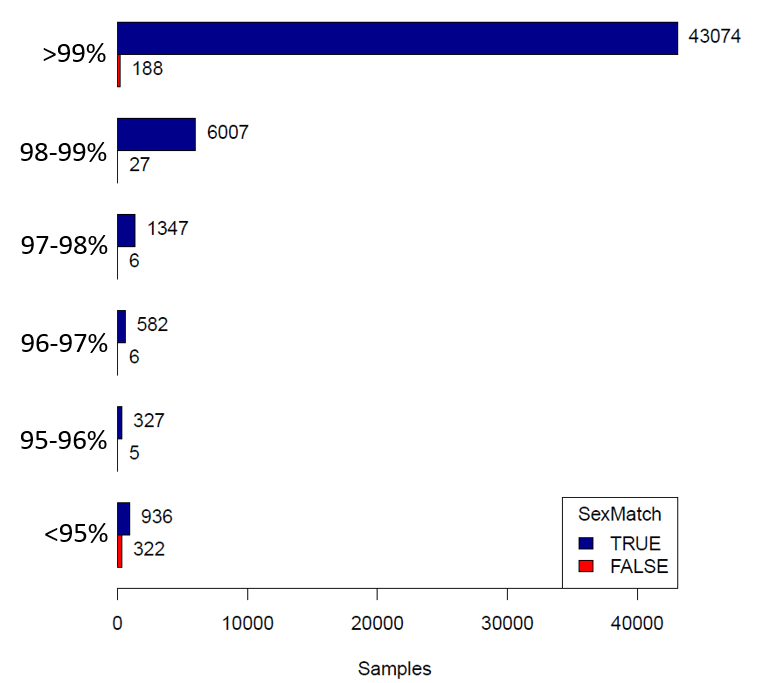

Supplementary Figure 3. Performance of genotyping stratified on call rates and sex match. Sample QC was primarily based on call rates, requiring more than 95% for a pass. A secondary validation was done using genotype sex, requiring that they match the electronic records. A total of 52,827 samples were genotyped. 51,337 (97.2%) samples passed both call rate and sex checks. 1,258 (2.4%)samples failed call regardless of the sex estimate matched. 232 (0.4%) samples passed call rate checks but failed the sex check, of these 123 (0.2%) were re-included as the error in sex estimation could be explained. The remaining 109 samples with errors in sex estimates were removed from the study.

|  |  |
| --- | --- |
| Has changed CPR | 108 |
| Wrong sample | 107 |
| Aneuploidy | 10 |
| All other | 7 |

Supplementary Figure 4. Breakdown of the reasons that genotype sex estimates does not match that expected from the birth registry. The primary reason is that the CPR number has changed during the follow-up period in the study. As we coded sex ambiguous for persons who had changed the CPR, any call will results in a failed test regardless of what is recorded in either record. Another main contributor was that the wrong sample was retrieved from the biobank. The reason that the wrong sample is retrieved is primarily database errors in the biobank and reduced readability of labels resulting from decades of storage. We included all subject in the study if the reason for initial flagging did not lead us to question the correct sample was retrieved (i.e. all groups except the wrong sample).

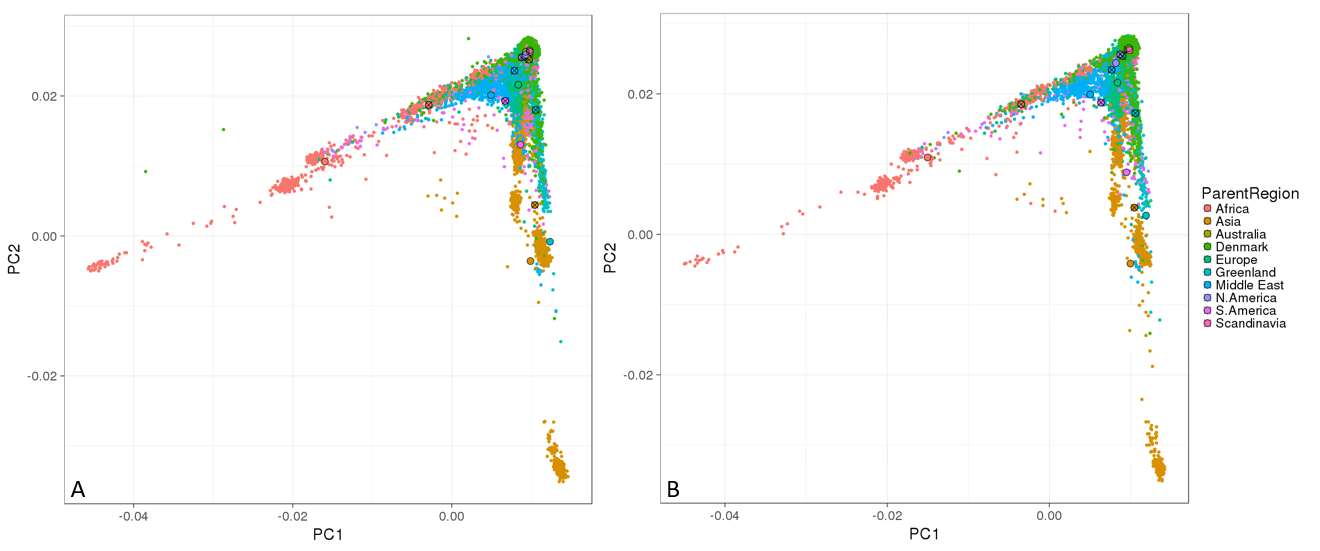

Supplementary Figure 5. PCA of the ﬁrst two principal components coloured according to the parental region of birth. Values are calculated for iPSYCH2012 (tile A, n=78,490) and iPSYCH2015i (tile B, n=51,460). Circles colour indicate mean values for the given parental group. Crossed Circles indicates both parents born abroad within the region indicated by the colour. Absence of cross indicates one Danish-born parent and one parent born in the region indicated by the colour.
